# Early Detection of CAR-T-Associated Neurotoxicity via Cytokine Monitoring in Serum

**DOI:** 10.64898/2026.03.03.26347491

**Authors:** Amit Parizat, Onit Alalouf, Danielle Sapir, Nivin Shibli, Ruth Perets, Dvir Aran, Ofrat Beyar Katz, Yoav Shechtman

## Abstract

Immune effector cell-associated neurotoxicity syndrome (ICANS) is a common and life-threatening complication of chimeric antigen receptor (CAR) T-cell therapy, with early detection being critical for timely intervention and improved outcomes. Cytokines such as interleukin-6 (IL-6) are key mediators of the inflammatory cascade underlying ICANS pathogenesis, but prospective clinical evidence for their predictive value is limited. Here we quantify IL-6 levels in a prospective cohort of 40 CAR-T patients (270 serum samples), using a simple in-house microfluidic bead immunoassay. IL-6 levels measured by our assay were significantly associated with ICANS onset. Specifically, each ∼3.4-fold increase in IL-6 levels was linked to a 74% increase in the odds of developing ICANS the following day, independent of other clinical variables. Overall, we show the prognostic value of IL-6 for next-day ICANS, demonstrate the potential of frequent cytokine measurement to guide CAR-T patient management, and develop a simple experimental method to perform such monitoring.

---

Chimeric antigen receptor (CAR) T-cell therapy has revolutionized the treatment of hematological malignancies such as B-cell lymphomas and leukemias. By engineering the patient’s T-cells to recognize and attack cancer cells, CAR-T has achieved impressive remission rates in cases where other treatments have failed^1^. However, these therapies are also associated with severe immune-related toxicities, particularly cytokine release syndrome (CRS) and immune effector cell-associated neurotoxicity syndrome (ICANS)^2,3^.

While CRS is often manageable, ICANS remains one of the most severe and unpredictable complications of CAR-T therapy. It occurs in approximately 20–60% of patients, depending on the CAR-T product and patient population, and typically emerges between 3 to 9 days after infusion, although cases have been reported even weeks or months later, depending on the CAR-T product used^1,2,4–6^. ICANS can present with symptoms ranging from mild confusion and language disturbances to seizures, cerebral edema, and coma, and in some cases can be fatal^2^.

Timely identification of patients at risk for ICANS is crucial to enable early therapeutic interventions, such as corticosteroids or anti-IL-6 therapies, which may mitigate neurotoxicity and improve outcomes^2,3,7–10^. However, current practice hinges on neurological assessment (e.g., ASTCT ICE scoring) after symptoms appear, constraining opportunities to intervene early^3,11,12^. Thus, there is a critical unmet need for biomarkers that reveal immune dysregulation before overt neurotoxicity.

Among such biomarkers, cytokines including IL-6, IL-15, IL-10, GM-CSF, and IL-1β are commonly elevated in ICANS and have been linked to its onset and severity^2,7,8,13^. Specifically, IL-6 plays a central role in the inflammatory cascade and is the target of the FDA-approved drug tocilizumab, making it a high-priority candidate for clinical monitoring. Retrospective studies have associated elevated IL-6 with ICANS onset, but prospective evidence demonstrating its predictive value is lacking. This gap largely reflects technological limitations – routine cytokine surveillance is uncommon because most assays are batch-based rather than random-access, too centralized, costly, and time-consuming for daily single-sample use, while others lack the sensitivity to detect clinically meaningful fluctuations^7,8,13–15^. For example, ELISA and multiplex bead panels (Luminex/MSD) are batch-oriented with multi-hour workflows^16^; ultra-sensitive digital immunoassays (e.g., Simoa) and centralized clinical ECLIA analyzers (e.g., Cobas) are expensive and impractical for on-demand single-sample runs^17,18^; and lateral-flow strips lack the precision required for serial IL-6 monitoring in this context^19–22^.

Accordingly, continuous monitoring requires a random-access, single-sample assay that can be run on demand from small serum volumes using standard laboratory infrastructure. The platform should be simple, affordable, and reproducible to support mechanistic studies, yet sufficiently precise across IL-6 concentrations relevant to ICANS, with short turnaround and minimal hands-on time. In this context, reliable quantification suitable for serial measurements is highly important, whereas extremely high analytical sensitivity is less relevant.

Here we show that routine cytokine monitoring in a standard laboratory setting could enable earlier identification of patients at risk for CAR-T-associated neurotoxicity. To achieve this, we developed a custom microfluidic bead immunoassay that combines ELISA-level accuracy with short turnaround time, minimal sample preparation, and random-access single-sample operation. This approach enabled daily cytokine tracking throughout CAR-T treatment and revealed a temporal association between IL-6 elevation and ICANS onset. Beyond this study, the assay establishes a technical foundation for future random-access, point-of-care platforms enabling individualized cytokine surveillance during cellular immunotherapies.

## Results

### Prospective monitoring of IL-6 dynamics during CAR-T treatment

To assess whether IL-6 dynamics reflect immune activation associated with neurotoxicity, we prospectively enrolled 40 patients with relapsed or refractory B-cell lymphoma undergoing CAR-T therapy between May 2023 and March 2025. Clinical characteristics are presented in **Table S1** and summarized in **Table 1**. Patient demographics showed an equal gender distribution, with most patients aged 60-80 years. Most received Yescarta (87.5%) as their CAR-T product, with smaller subsets receiving Kymriah (7.5%) or Tecartus (5%). CRS and ICANS were diagnosed and graded according to the ASTCT consensus grading system^3^. The average time to CRS onset was 1.9 days post-infusion (**Fig. S1A**), primarily manifesting as grade 1 (47.5%) or grade 2 (30%) toxicity. ICANS appeared later, with a mean onset of 5.4 days post-infusion (**Fig. S1B**), predominantly as grade 1 (17.5%) or 2 (12.5%).

**Table 1:** Patient clinical characteristics.

|  |  |  |  |  |
| --- | --- | --- | --- | --- |
| Number of patients | 40 |  |  |  |
| Median age at infusion (SD), range | 66 (±13), 23-80 |  |  |  |
| Gender distribution, % | 50 Female, 50 Male |  |  |  |
| CRS grade (mean onset day: 1.9) | 1 | 2 | 3 | 4 |
| n (%) | 19 (47.5) | 12 (30) | 6 (15) | 1 (2.5) |
| ICANS grade (mean onset day: 5.4) | 1 | 2 | 3 | 4 |
| n (%) | 7 (17.5) | 5 (12.5) | 3 (7.5) | 3 (7.5) |
| CAR-T product | Yescarta | Kymriah | Tecartus |  |
| n (%) | 35 (87.5) | 3 (7.5) | 2 (5) |  |

In addition to routine blood tests, daily morning serum samples were collected from day -8 to day +14 relative to the CAR-T infusion, with occasional missing days due to sample availability limitations (full collection data in **Table S1**). The collected samples were analyzed for IL-6 concentrations using a microfluidic bead immunoassay developed for this study and validated against the clinical gold-standard Cobas analyzer (**Fig. 1**).

**Figure 1:**
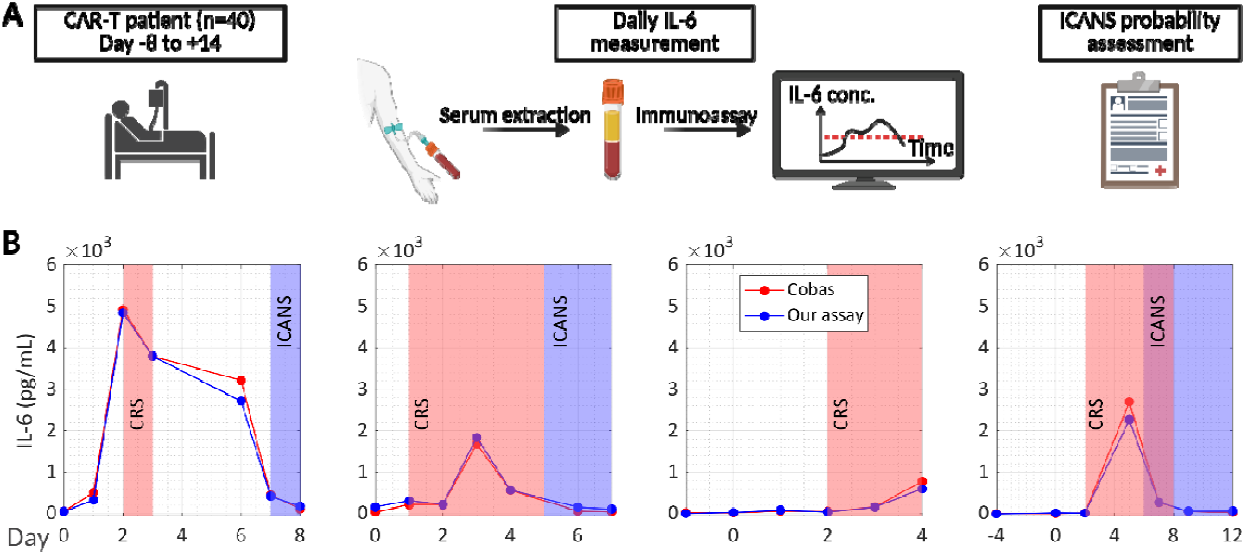
Daily IL-6 monitoring in CAR-T patients. **A**. Sampling and analysis workflow. **B**. Longitudinal serum IL-6 measurements in four representative patients. IL-6 was quantified with the microfluidic bead immunoassay developed for this study (blue) and a Cobas analyzer (red). Periods of CRS and ICANS are indicated by red and blue shaded regions, respectively.

### Development and validation of a microfluidic bead immunoassay for cytokine quantification

We developed a microfluidic bead immunoassay for quantitative IL-6 detection in serum. The assay uses a sandwich immunoassay on magnetic beads and reads out fluorescence while beads flow through a microfluidic channel (**Fig. 2A**). Dual illumination – laser for fluorescence and a temporally modulated LED in dark field for bead scattering – enables single-channel co-localization and robust bead identification during flow (**Fig. 2B-C**). Automated image analysis aggregates per-bead fluorescence to a sample-level signal, and absolute concentrations are obtained from a two-point standard curve. After 6 minutes of acquisition, hundreds of beads are analyzed per sample (see **Fig. S2** for the effect of acquisition duration on assay sensitivity), supporting on-demand, single-sample operation. The assay achieved a limit of detection of 72.61 pg/mL for IL-6 (**Fig. 2D**) and demonstrated linearity, precision, reproducibility, and specificity suitable for clinical monitoring (**Fig. S3**). For future applications such as multi-analyte monitoring, the method also allows simple multiplexing, by including beads with different sizes. (**Fig. S4**).

**Figure 2:**
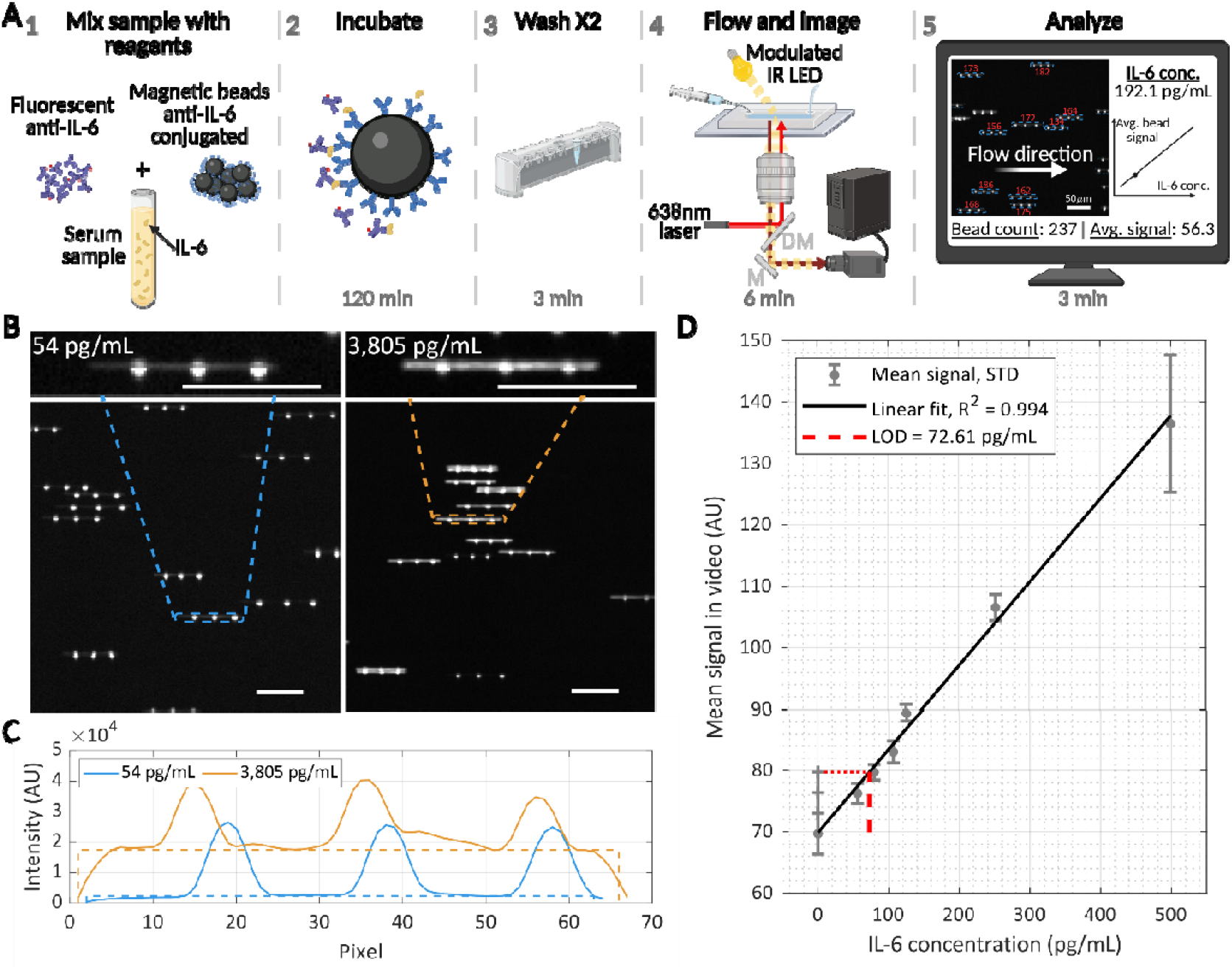
IL-6 quantification in serum using the microfluidic bead immunoassay. **A**. Schematic of the assay workflow. (1) Serum is mixed with magnetic beads conjugated to capture antibodies and with fluorescently labeled detection antibodies. (2) During incubation, IL-6 molecules form sandwich complexes on the beads. (3) Beads are washed twice using a magnetic rack. (4) The sample is flowed through a microfluidic channel and are imaged using a laser and a temporally modulated LED for bead localization (M – mirror, DM – dichroic mirror, IR – infrared). (5) Images are analyzed to localize beads, quantify fluorescence, and infer IL-6 concentration. **B**. Representative video frames showing flowing beads incubated with serum containing low (∼54 pg/mL, left) and high (∼3800 pg/mL, right) IL-6 concentrations. Insets show zoomed-in regions with beads – continuous lines are fluorescent bead streaks during exposure; the dot triplets result from scattering from pulsed LED illumination as the bead flows (scale bars - 50 μm). **C**. Intensity profile of the horizontal projection of the beads presented in the insets of panel B, with matching colors. Dashed lines represent the height and width of the fluorescent signal in each bead, determined algorithmically (see Methods section). Their multiplication is the bead signal. **D**. The LOD of IL-6 in human serum was determined by a linear fit (R^2^ = 0.994) of 7 samples of up to 500 pg/mL (three replicates each; eight replicates for blank serum). Horizontal dashed red line is the average intensity of a blank sample + 3×SD. Vertical dashed red line is the intersection with the fitted linear curve, determining a LOD of 72.61 pg/mL.

Across 270 samples, IL-6 values spanned approximately 73–15,000 pg/mL and closely matched those obtained with the Cobas analyzer over the dynamic range (**Fig. S5**). The median difference between Cobas and our assay measurements was -5.94 pg/mL, with a standard deviation of 189 pg/mL excluding four outliers exceeding 11,000 pg/mL. This degree of variation is consistent with previously reported inter-platform variability in cytokine quantification, where absolute concentrations can vary substantially between commercial assay systems despite strong correlations^16,17,23,24^.

### Association between elevated IL-6 levels and ICANS onset in CAR-T patients

Using our bead assay, we tested IL-6 as an ICANS predictor and compared its performance to other clinical data associated with this neurotoxicity^8,13^, such as CRS onset, day from CAR-T infusion, age, platelet counts, and CRP and ferritin levels. Following CAR-T infusion, patients exhibited characteristic toxicity patterns. CRS manifestations emerged in most patients soon after treatment, peaking between days 3-5, and resolving by day 10 (**Fig. 3A**). ICANS demonstrated a more variable timing of onset, occurring in fewer patients and appearing at different timepoints across the post-infusion period, with a mean onset of 5.4 days but ranging from as early as day 1 to as late as day 9. ICANS was most abundant around days 7-10, and when present, showed a longer resolution phase than CRS, often extending beyond day 14 (**Fig. 3A**). The kinetics of these clinical toxicities were paralleled by changes in serum IL-6 levels. Mean IL-6 concentrations remained relatively stable during days 0-1, began rising on day 2, and peaked at day 5, followed by a rapid decline to near-baseline by day 7 (**Fig. 3A**). Notably, the IL-6 elevation preceded and followed the development of both CRS and ICANS, but with different temporal relationships to each syndrome. When stratifying IL-6 levels by CRS grade, excluding pre-infusion days (**Fig. 3B**), we observed a clear positive correlation, with median IL-6 values progressively increasing from ∼72 for grade 0 (i.e. no CRS) to ∼8200 pg/mL for grade 4 (*p* = 3×10^-12^). Notably, no such grade-dependent relationship was evident for ICANS, suggesting different pathophysiological mechanisms.

**Figure 3:**
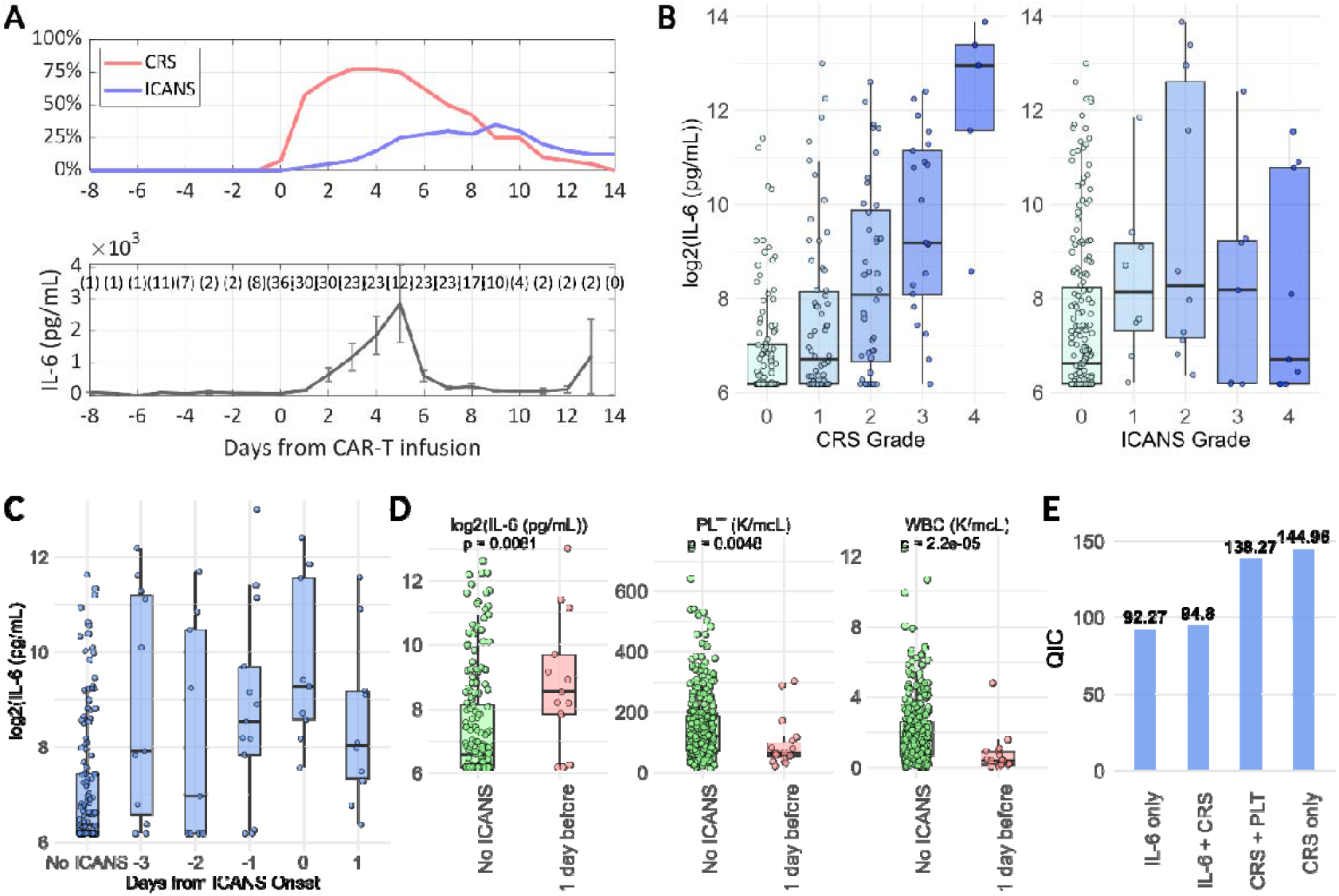
IL-6 monitoring for assessing correlation to ICANS onset. **A**. Percentage of patients with CRS and ICANS over time. Below, IL-6 levels over time as measured by BMF, averaged across all patients. Numbers in brackets indicate the number of samples per time point. Error bars represent the standard error (SDT/√n). **B**. Distribution of IL-6 levels in post-infusion days according to CRS/ICANS levels. **C**. Distribution of IL-6 levels by days to ICANS onset (any ICANS grade). **D**. Distribution of IL-6, platelets (PLT), and white blood cell (WBC) levels, one day before ICANS onset compared to days with no ICANS (post-infusion days, not including post-ICANS onset values). P-values were determined using Wilcoxon test. **E**. Comparison of QIC values across the GEE models. Lower QIC values indicate better model performance.

To investigate the potential of IL-6 as a predictive biomarker for ICANS development, we analyzed the temporal relationship between IL-6 elevation and subsequent ICANS onset. We observed that IL-6 levels were significantly elevated one day before ICANS diagnosis, compared to patients who did not develop ICANS (**Fig. 3C-D**). This elevation was further pronounced on the day of ICANS diagnosis, followed by a decline on the subsequent day, likely reflecting therapeutic interventions. When dichotomizing outcomes based on ICANS development the following day (**Fig. 3D**), we identified significantly higher IL-6 levels in patients who subsequently developed ICANS (*p* = 0.008). Additionally, lower platelet (PLT) counts and white blood cell (WBC) counts were inversely associated with impending ICANS (*p* = 0.005 and *p* < 0.0001, respectively).

To account for potential confounding factors, we constructed a series of generalized estimating equation (GEE) models with an autoregressive correlation structure to evaluate the association between IL-6 levels and the development of ICANS on the following day (**Fig. 3E**). In the base model adjusting only for IL-6, higher IL-6 levels were significantly associated with increased odds of ICANS (odds ratio (OR) 1.86, 95% confidence interval (CI) [1.22, 2.84], *p* = 0.004). A separate model that included only CRS also showed a significant association (OR 1.95, 95% CI [1.29, 2.97], *p* = 0.002; **Fig. S1C**), indicating that both variables are individually informative. When IL-6 was added to the CRS-only model, only IL-6 remained significant (IL-6 OR 1.74, 95% CI [1.04, 2.92], *p* = 0.034), suggesting that IL-6 provides predictive value beyond CRS alone. Notably, a one standard deviation increase in log_2_(IL-6), equivalent to ∼3.4-fold increase in IL-6 concentration, was associated with a 74% increase in the odds of developing ICANS, independent of CRS grade. A similar model including PLT and CRS did not yield a significant association between ICANS and PLT.

To compare candidate GEE models while guarding against overfitting, we compared the quasi-likelihood under the independence model criterion (QIC) values of all four models (**Fig. 3E**). QIC, an extension of the Akaike information criterion (AIC) for GEE models, balances model fit and complexity in the context of correlated data, with lower QIC values indicating better model performance^25^. The model incorporating IL-6 alone demonstrated the best model fit, achieving the lowest QIC score. The observed increase in QIC upon adding CRS to the IL-6 model reflects that CRS does not contribute substantial independent predictive information beyond IL-6 and instead unnecessarily increases model complexity, which may cause overfitting. These findings underscore the robust and consistent predictive value of IL-6 across all models, highlighting its potential as a reliable biomarker for anticipating ICANS development even when accounting for other clinical indicators.

## Discussion

In this prospective cohort, we developed an assay suitable for on-demand, daily cytokine monitoring, and used it to show a temporal association between IL-6 levels and subsequent ICANS onset. IL-6 levels were higher on the day before ICANS diagnosis and remained elevated at onset. In GEE models, a one standard deviation increase in log2(IL-6) (≈3.4-fold) was associated with 74% higher odds of next-day ICANS, independent of CRS grade. These results extend prior retrospective observations by demonstrating a prospectively observed, temporally ordered relationship between IL-6 elevations and short-term ICANS risk.

Our findings support incorporating routine IL-6 monitoring into early-warning workflows for ICANS, enabling timelier clinical evaluation and intervention. Because ICANS often emerges within the first week after infusion, a simple, same-day IL-6 readout can complement neurologic assessments and standard labs to focus attention on patients with rising short-term risk. Early identification may also permit earlier therapeutic actions (e.g., corticosteroids or anti-IL-6 therapy) before overt neurotoxicity, which is particularly relevant in outpatient care pathways where rapid escalation can be more challenging.

From an implementation standpoint, serial quantification is central. The microfluidic bead immunoassay enabled on-demand, single-sample runs from small serum volumes and tracked a clinical analyzer across the relevant dynamic range, supporting practical daily surveillance in standard laboratories. Such surveillance could complement neurologic assessments and routine labs to facilitate earlier clinical evaluation and treatment during the peri-infusion window when ICANS risk is highest.

In centers already obtaining morning labs after CAR-T infusion, same-day IL-6 results could inform thresholds for closer monitoring or pre-emptive therapy in patients with rising values, particularly in outpatient care. While IL-6 alone showed consistent association in this study, future work should test whether combining IL-6 with commonly available measures (e.g., CRP, ferritin, platelets) improves short-term risk stratification without the complexity of broad cytokine panels.

Although the assay supports random-access single-sample runs, this study processed stored aliquots and did not record operational timelines. Phlebotomy-to-result turnaround, hands-on time, run failure rate, and inter-operator reproducibility were not quantified. Therefore, operational validation is needed to substantiate the timeliness of early detection. Pragmatic pilot studies should assess whether same-day IL-6 results can be routinely delivered in time to inform clinical decisions.

This single-center design and product distribution (predominantly Yescarta) motivates external validation across CAR-T products and practice settings. Furthermore, once-daily sampling may miss finer dynamics around onset or interventions between draws, potentially attenuating associations, and clinical workflows and treatment thresholds may differ across institutions. Multi-site studies should (i) verify the next-day association and define clinically actionable IL-6 thresholds, (ii) assess whether responses to rising IL-6 (e.g., earlier evaluation, corticosteroids, or anti-cytokine therapy) reduce ICANS severity or resource use, and (iii) evaluate higher-frequency or continuous sampling to refine lead-time estimates and enable dynamic risk-estimation updating models. In parallel, selectively extending panels, potentially compatible with our assay’s multiplexing capabilities, may be informative in specific scenarios. For example, IL-1β^26,27^ and IL-15^28^ for tighter inflammatory profiling^26^; TF and PECAM-1 for CRS-related coagulopathy^29^; and CRP, ferritin, and IL-6 for early immune-effector cell-associated hematotoxicity^30^ (e.g., guiding G-CSF decisions).

Prospectively observed IL-6 elevations precede ICANS onset and are associated with increased short-term risk, supporting incorporation of routine IL-6 monitoring into CAR-T care pathways to enable earlier clinical decision-making. The assay’s agreement with a clinical analyzer and single-sample operation suggests near-term deployability in standard laboratory settings, while larger validation studies can establish thresholds and protocols for proactive neurotoxicity management.

## Methods

### Participants

All clinical studies were approved by the Israeli Ministry of Health Ethics Committees (Helsinki application RMB-0636-22). Informed consent was obtained from all individuals before blood sampling.

### Serum collection

Blood samples were collected in Vacuette CAT serum separator clot activator tubes (item number 456005). Blood immediately was centrifuged (10 min at 1,500 RPM and 5 °C), and the supernatant was transferred to a 15-ml tube. Serum was then aliquoted in low-bind 0.2 ml tubes and stored at −80°C for analysis.

### Method validation

We compared our immunoassay for IL-6 detection with the clinical gold-standard Cobas® e 411 analyzer and Elecsys IL-6 kit (Roche) for all serum samples. The samples were thawed and vortexed. They were not diluted unless IL-6 values exceeded 5000 pg/ml, in which case measurements were repeated with a 1 in 5 dilution.

### Bead preparation

Antibodies (R&D systems, MAB206 or Abcam, ab241807 (capture antibody)) were coupled to magnetic beads according to manufacturer instructions. Before coupling, the antibody solution was buffer-exchanged using an ultrafiltration vial (Biotium, 22004, MWCO=10k); an equal volume of MES buffer (concentration depending on the bead type, see below) was added to the vial with the antibody solution, the vial was centrifuged for 1.5-3 min at 11,700xg, and the antibody was recovered with MES buffer to the required concentration.

The magnetic beads were thoroughly resuspended in the original vial for ½ hr using a roller mixer, and the required amount for coupling was transferred to a low bind tube.

#### Chemicals

MES (99%, MW=195.2, Acros Organic, 274140500); PBS (Sigma, p5368); Tween-20 (Sigma, P-9416); BSA (Sigma, A7906); EDAC hydrochloride (Sigma, 341006). Solutions were filtered with 0.2 µm filter system or Millex – GV 0.2 µm.

#### Coupling antibodies to Dynabeads® M-270 Carboxylic Acid magnetic beads (Invitrogen, 14305D)

the beads were washed twice with 0.01 M NaOH and then 3 times with deionized water by incubating them for 10 min in the roller mixer and recovering them using a magnet. EDAC solution (62 mg/ml) was prepared in deionized water just before usage. The EDAC solution (double the original volume) was added to the recovered beads and vortexed until all the beads were in solution. The beads were incubated for 30 min at room temperature in the roller mixer. The beads were recovered from the EDAC solution and washed once with double volume of cold, deionized water and once with 50 mM MES, pH 5, as quickly as possible, to avoid hydrolysis of the activated carboxylic acid groups. The pre-prepared antibody in 50 mM MES, pH 5, was added and the suspension was incubated for 30 min at room temperature. Antibody concentration in the supernatant was measured to determine binding efficiency using the Implen-NP80 spectrophotometer. The beads were washed 4 times with 50 mM Tris buffer, pH 7.4 by incubating in the roller mixer for 3 minutes and were then washed 4 times with PBS, 0.1% Tween, 0.1% BSA buffer and finally resuspended in the original volume in PBS, 0.1% Tween, 0.1% BSA, 3 mM sodium azide buffer and stored at 4^°^C.

#### Coupling antibodies to Dynabeads® MyOne™ Carboxylic Acid magnetic beads (Invitrogen, 65011)

the beads were washed twice with an original volume of 15 mM MES buffer pH 6.0 and resuspended in 1/10th of the original volume in 15 mM MES, pH 6.0. An equal volume of 10 mg/ml EDAC (made just before usage in cold deionized water) was added to the beads and incubated on the roller for 30 minutes at room temperature. The pre-prepared antibody in 15 mM MES, pH 6, was added to the beads and the tube was incubated in the roller mixer overnight at room temperature. Antibody concentration in the supernatant was measured to determine binding efficiency using the Implen-NP80 spectrophotometer. The beads were washed twice with a PBS/0.1% Tween-20 buffer in the roller mixer for 10 minutes and resuspended to the original volume in PBS/0.1% Tween-20/0.1% BSA/3 mM sodium azide buffer and stored at 4^°^C.

For the 2.8 μm beads, before experimental usage, the beads were diluted (1 in 50) in PBS with 4% BSA and photobleached by concentrating the beads with a magnet and placing the tube in front of a 2W 638 nm laser beam with 0.5 ND filter for 4 hours (see supplementary **Fig. S6** for photobleaching optimization), during which the beads were mixed and concentrated every hour to ensure optimal photobleaching of all beads.

### Antibody labeling

Detection antibody (R&D Systems, AF-206-NA or Abcam, ab241807 (detection antibody)) was labeled using DyLight® 633 antibody-conjugation kit (abcam, ab201802) according to the manufacturer’s protocol, and stored at 4°C.

### Immunoassay

50 μL of CAR-T patient serum samples (thawed on ice) or spiked serum samples (IL-6 (R&D systems, 206-IL) in commercial human serum (Capricorn Scientific, HUM-3B)) were diluted 1 in 5 in a dilution buffer (PBS, 0.05% Tween20, 7mM EDTA), photobleached bead solution (about 400,000 beads), and 0.2 μg/mL detection antibody, and incubated together at 25°C in the dark, 1000 RPM shaking. After 2hr incubation in a tube, the samples were washed twice using a magnetic rack and wash buffer (PBS, 0.05% Tween20, 5mM EDTA) to a final volume of 100 μL. The tube was then connected to the flowing system for IL-6 detection and measurement.

For multiplexed measurement, 50 μL of spiked serum samples were diluted 1 in 5 in a dilution buffer, photobleached IL-6 bead solution, IL-1β bead solution, and 0.2 μg/mL of each detection antibody. The rest of the assay was identical.

For patient sample measurements, 2 samples of patients with known concentrations, measured with the Cobas analyzer were used to obtain a linear calibration curve. The commercial human serum was tested using Cobas and has baseline IL-6 levels of <1.5 pg/mL.

Experiments in which less than 100 beads were measured were discarded to avoid high sampling error. This happened rarely, due to blockage of the microfluidic channel. For each sample, an average of 238 beads (±93) was imaged.

### Microfluidics setup

The flow was driven by an air pressure controller (Fluigent Push-Pull, Fluigent Flow EZ). A 20 μm height microfluidic channel was used (ChipShop, Fluidic 180) with 1/32” inner diameter Tygon® silicone tubing (Cole-Parmer).

### Optical setup and image acquisition

Imaging was performed using an inverted microscope (Nikon, Eclipse Ti2-E) with a x40/0.75 NA air objective (Nikon, CFI Plan Fluor 40X) and custom-made filter cube containing dichroic mirror (Semrock, Di03-R635-t3-25x36) and long pass emission filters (Semrock, BLP01-635R-25, Chroma, ET665lp). A high power 2W 638 nm laser with a ND 0.5 filter (Thorlabs, ND05A) was coupled with a multimode optic fiber (Thorlabs, M103L05) and connected to the microscope. A broadband 850 nm LED (30 nm FWHM, Thorlabs, M850L3) with LED Driver (Thorlabs, LEDD1B) was collimated using an aspheric condenser lens (Thorlabs, ACL2520U-B), and filtered using 850nm LPF (Thorlabs, FELH0850). The LED was positioned in a sharp angle relative to the objective lens for dark field illumination. To modulate the light source, a custom-made chopper (3D printed) located in front of the LED was rotated using an electric engine (Pololu Robotics and Electronics) and a power source. A field of view (FOV) of 330 μm × 330 μm was recorded using a sCMOS camera (Teledyne photometrics, Prime 95B) with 16 bit-depth, 0.4s exposure time and 2 by 2 binning. The flow was imaged for 6 minutes, obtaining 900 frames.

### Video analysis

Image analysis was done using ImageJ, Python (Visual Studio Code) and Matlab R2024b.

#### Preprocessing and noise reduction

To reduce the data size for faster analysis, further 2 by 2 binning was done to obtain frames of 300 by 300 pixels. In addition, a semi-constant background was reduced by subtracting the median of 10 frames around each frame. Frames with outlier mean value (higher or lower than 4 standard deviations from the mean of the frame means in the video) or with more than 30 localizations of beads were discarded. Due to LED modulation, an additional horizontal wave-like noise was added. To reduce it, a smoothing of the horizontal projection of each frame (after thresholding to remove bead’s signal) was also subtracted from it.

#### Bead localization and fluorescent intensity calculation

To locate the flowing beads in each frame, the YOLO11 algorithm (Ultralitics^31^) was trained and used on 8-bit RGB converted frames. Then, localizations near the edges of the frame were removed and the rest were vertically summed into 1D vectors for further analysis. Only localizations containing 3 or 4 sharp peaks were considered as beads and their fluorescent signal was calculated by multiplying the bead’s width by the median of the minima of the smoothed 1D vector (see supplementary **Fig. S7**). This value was divided by 1000 and by a normalization value for FOV dependence intensity correction (see supplementary **Fig. S8B**).

#### Post-processing

In each frame, in case of beads which were too close to each other, only a single bead with the highest localization certainty was left for final analysis. Beads in consecutive frames were identified and labeled, and each bead’s average intensity was calculated (4-5 frames). Beads with outlier fluorescent signals (more than 3.5 standard deviations above the average) were removed.

#### IL-6 concentration derivation

For each video, the mean value of fluorescent signal of detected beads was calculated and considered as the video’s signal. Using the video signal of the two standard samples, a linear fit was calculated and used to derive the IL-6 concentration of an unknown sample.

#### Statistical analysis

was done using RStudio. Before analysis, all IL-6 levels below the LOD (72.61 pg/mL) were considered as 72.61 pg/mL, and the log_2_ of all values was calculated. Outcome was defined by 0 if ICANS did not start in the next day, and 1 if it did. Days pre-infusion and after ICANS onset (including the day of onset) got NA outcome value, and later removed. p-value of the difference between IL-6 levels in different CRS grades was done using Kruskal-Wallis rank sum test (**Fig. 3B**). p-value of the differences between outcome 0 and outcome 1 days (**Fig. 3D**), was done using Wilcoxon test. In the generalized estimating equation (GEE) models, a binomial link function was used, and days with missing IL-6 or PLT values were excluded.

## Data Availability

All data produced in the present study are available upon reasonable request to the authors

## Acknowledgments

We would like to thank Anna Shur and Marielle Kaplan from the Biochemical Lab at Rambam hospital for performing the Cobas measurements and providing valuable advice, and Iris Halamish from the IT department at Rambam hospital for extracting the laboratory data for this study. We greatly appreciate the technological advice provided by Tal Gilboa from Wyss Institute, Harvard University, and Oded Lewinson and Nurit Livnat-Levanon from the Faculty of Medicine at the Technion. We also would like to acknowledge Ofri Goldenberg for conducting data analysis, and Tav Nahimov and Paz Keidar for their role in the experimental setup. Finally, we extend our deepest gratitude to the patients and the staff at Institute of Hematology and Bone Marrow Transplantation for their support and assistance in sample collection, which made this study possible.

