## Supplementary Information for "Early Detection of CAR-T-Associated Neurotoxicity via Cytokine Monitoring in Serum"

**Supplementary figures**

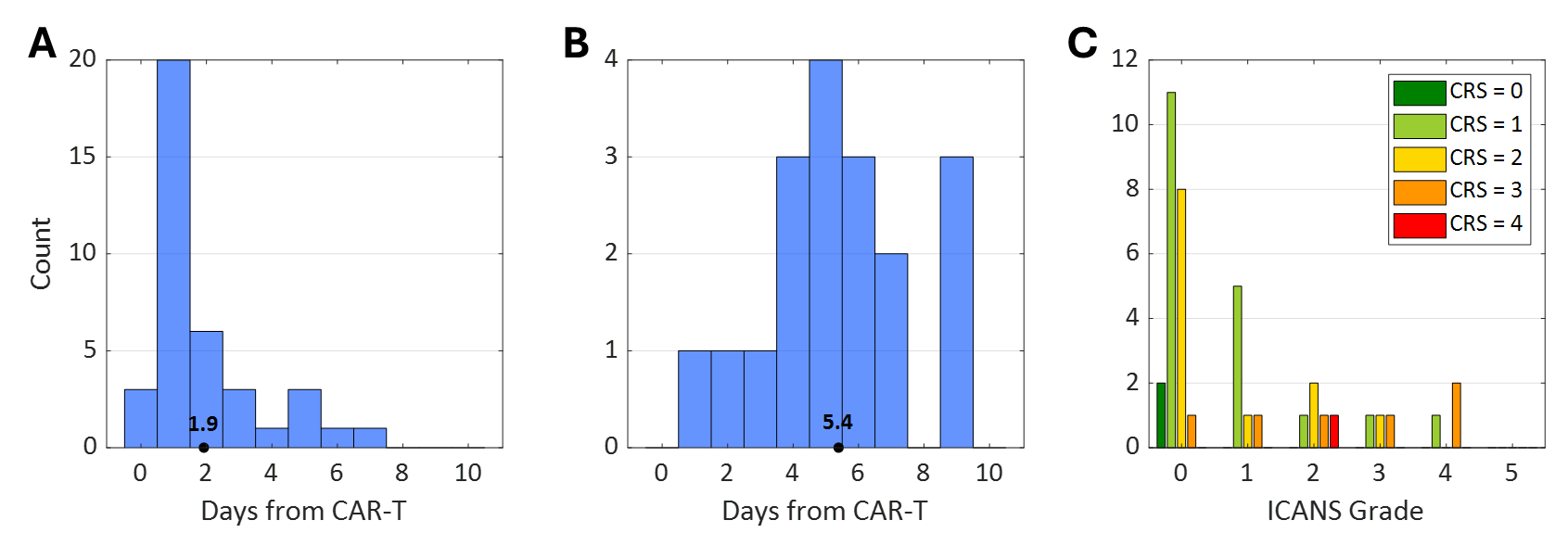

**Figure S1**: Patient characteristics related to CAR-T treatment-associated toxicities. **A.** Distribution of Cytokine Release Syndrome (CRS) onset days relative to CAR-T infusion. The black dot marks the mean onset day across patients. **B.** Distribution of Immune Effector Cell-Associated Neurotoxicity Syndrome (ICANS) onset days. The black dot marks the mean onset day. **C.** ICANS grade distribution stratified by CRS grade. A positive association is observed, with higher ICANS severity more frequent among patients experiencing higher CRS grades.

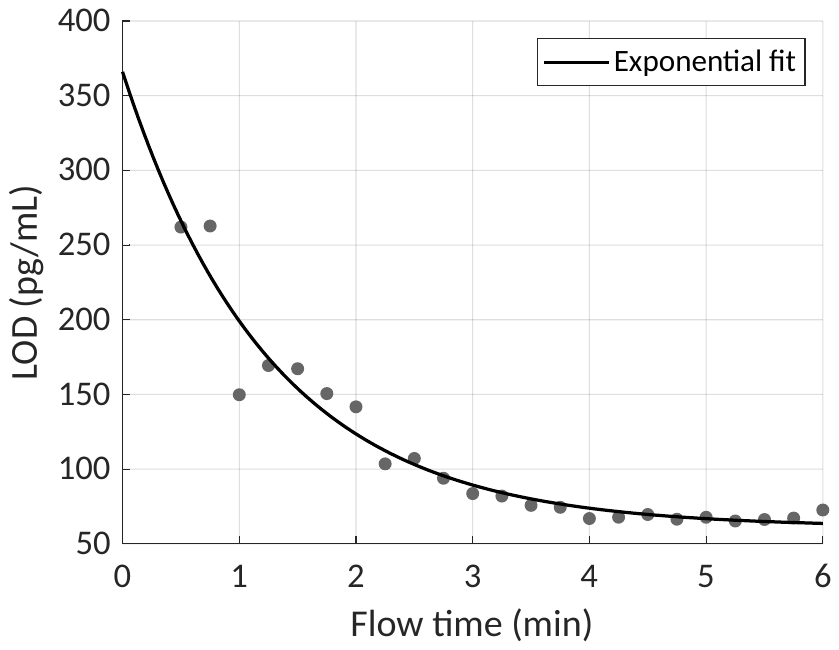

**Figure S2**: Limit of detection (LOD) as a function of sample flow time. The LOD was recalculated by progressively truncating video frames from the end of the recordings used in the LOD experiment shown in **Figure 1D**. LOD values were computed following the procedure detailed in the **Figure 1D** caption.

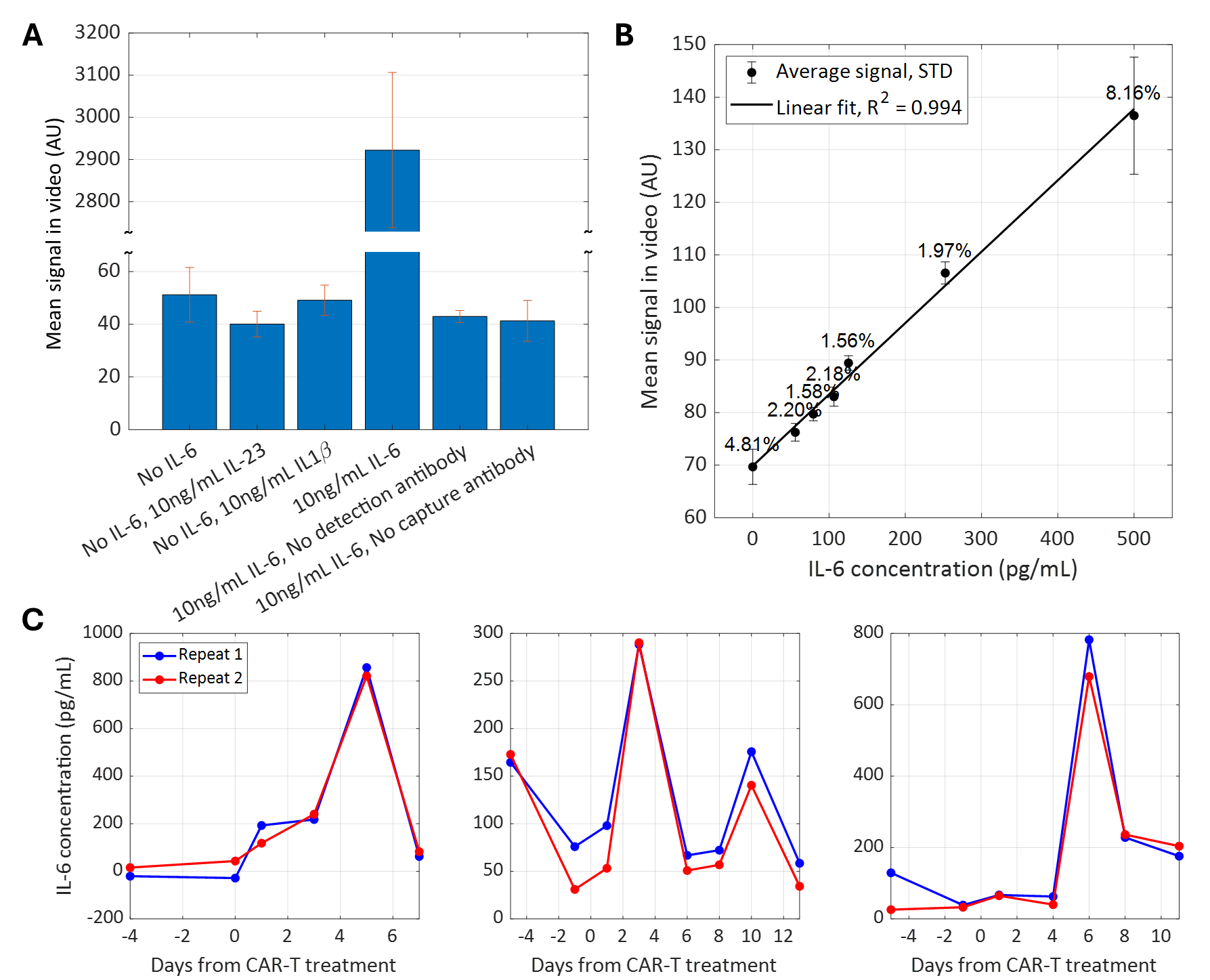

**Figure S3**: Biochemical characterization of BioMarkerFlow. **A.** Specificity control experiments for IL-6 detection. Signal measured in response to IL-6, IL-23, and IL-1β, and in negative control conditions lacking either the detection antibody or the capture antibody. Bars represent the standard deviation (STD) of three replicates. **B.** Intra-assay reproducibility (data taken from the LOD experiment shown in **Figure 1D)**. Coefficients of variation (%CV = 100% × STD / mean) are indicated above each bar. For the 0 pg/mL IL-6 sample, eight replicate measurements were used; for all other concentrations, three replicates were used. **C.** Inter-assay reproducibility using serum samples from three CAR-T-treated patients, measured on two different days with independent calibrations. Red and blue lines correspond to the two independent measurements per patient.

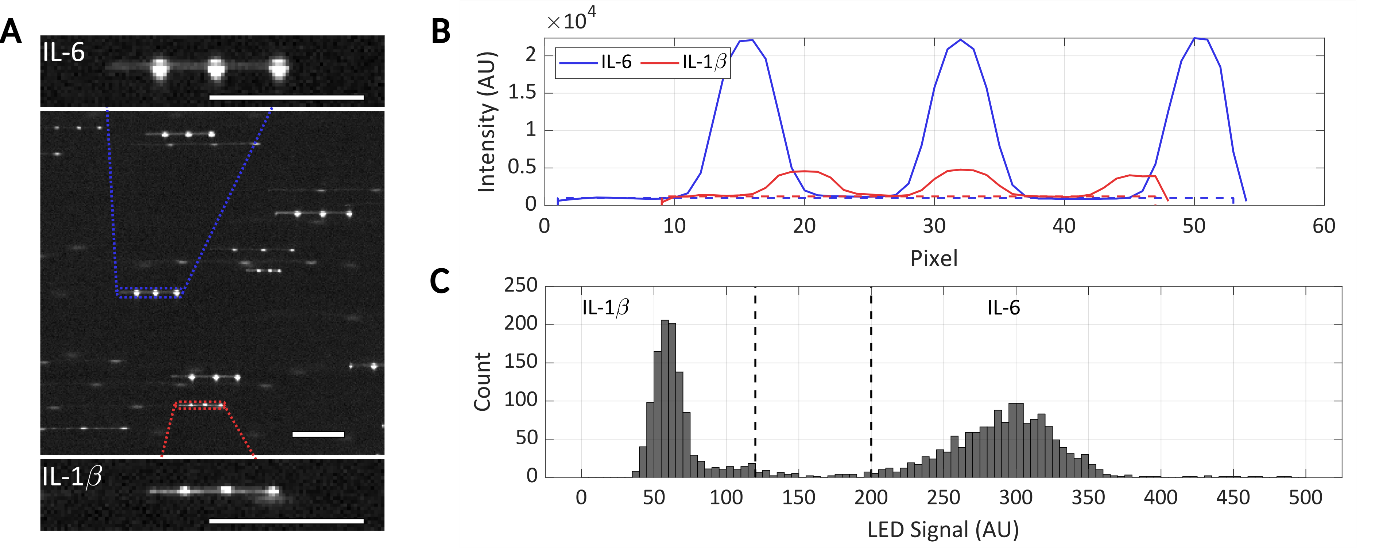

**Figure S4**: Proof of concept multiplexing capability. **A.** Representative frame from a multiplexed measurement of IL-6 and IL-1β. Upper inset shows a 2.8 μm bead targeting IL-6; lower inset shows a 1 μm bead targeting IL-1β (scale bars - 50 μm). **B.** Intensity profiles of the horizontal projection of the beads in panel A, with matching colors. Dashed lines represent the height and width of the fluorescent signal in each bead. Their multiplication is the bead signal. Since larger beads scatter more of the LED’s light, differentiation between beads reporting on each of the biomarkers was done based on the intensity of the signal in the peaks caused by the modulation. **C.** Histogram of LED signals for beads from the multiplexed measurement. LED signals are the area in F between the graph and the dashed horizontal line. Dashed lines define signal thresholds used to exclude ambiguous bead signals between the 1 μm and 2.8 μm bead populations, minimizing crosstalk.

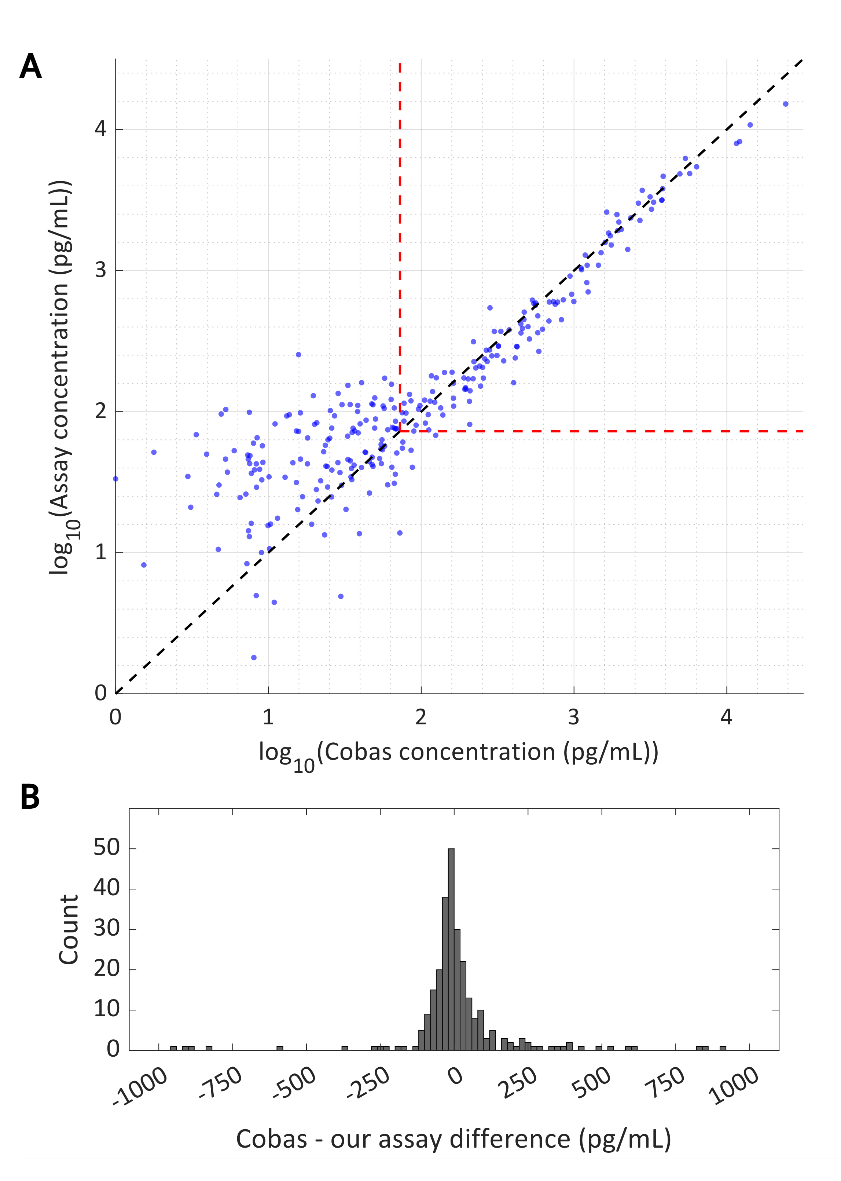

**Figure S5**: Our assay accurately measures IL-6 in CAR-T patients **A.** Comparison of IL-6 concentrations measured by our assay and Cobas analyzer in 270 serum samples from CAR-T patients. The black dashed line indicates perfect agreement (line of identity). Red dashed lines represent limit of detection of our assay. Concentrations are shown on a log₁₀ scale. **B.** Histogram of the differences between IL-6 concentrations measured by Cobas and our assay across the samples shown in panel A.

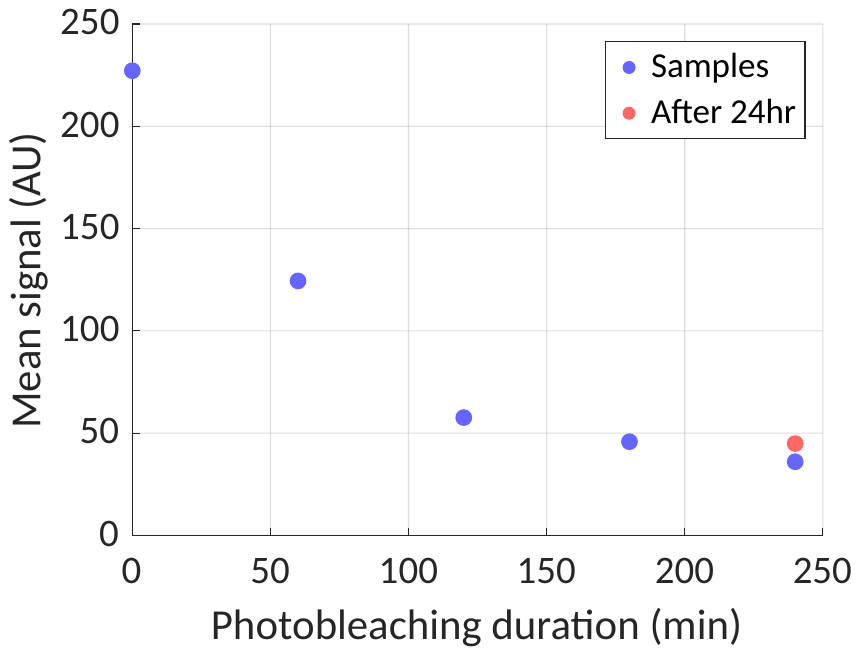

**Figure S6**: Bead photobleaching experiment for determining optimal photobleaching duration. Beads were diluted and subjected to varying durations of photobleaching, then immediately measured in the system (blue points). The orange point represents a sample that was photobleached for 240 minutes and measured after 24 hours, indicating signal stability over time. The mean signal (arbitrary units, AU) decreases with increased photobleaching time, approaching a stable baseline beyond 200 minutes.

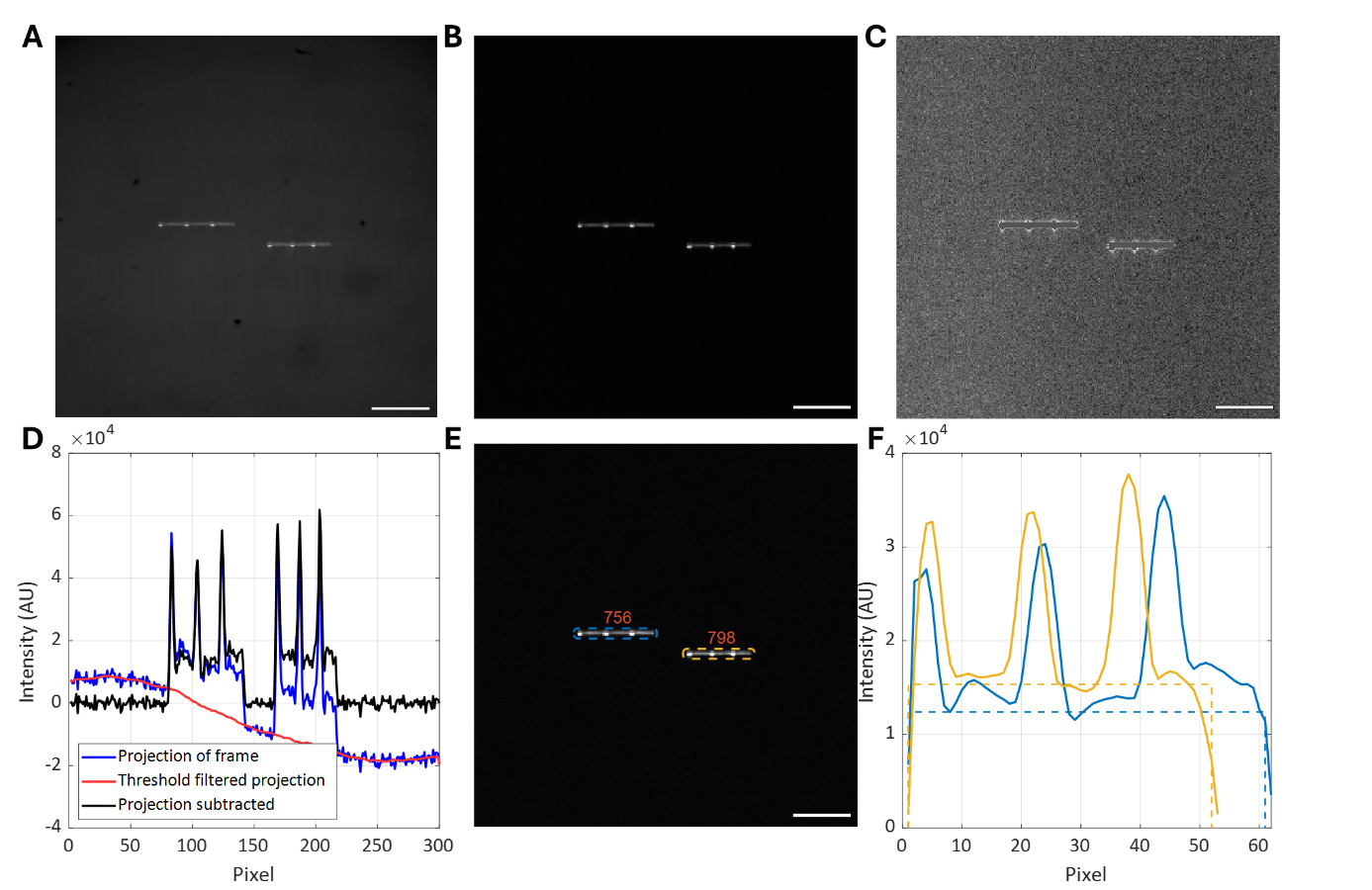

**Figure S7**: Visual summary of the video analysis pipeline for bead detection and fluorescent signal quantification. **A.** Raw frame (600 × 600 pixels) showing two fluorescent beads flowing through the microfluidic channel. **B.** Frame after semi-constant background subtraction and 2× binning (resulting in a 300 × 300 pixel image). **C.** Threshold-filtered version of panel B, isolating high-intensity regions corresponding to beads. **D.** Further background correction to eliminate horizontal wavy noise caused by LED modulation. The plot shows the horizontal intensity projection of panel B (blue), the smoothed thresholded projection from panel C (red), and their subtraction (black). **E.** Final processed frame showing localized beads, identified using the YOLO11 object detection algorithm. Fluorescence intensity values are labeled for each detected bead. **F.** Horizontal projections of the bead regions of interest (ROIs) shown in panel E, color-coded accordingly. The dashed horizontal lines represent the intensity values used as the height factor for each bead. The total fluorescence intensity is computed as the product of this height and the width of the bounding box, where the width is obtained from the YOLOv1 detection. All scale bars - 50 μm.

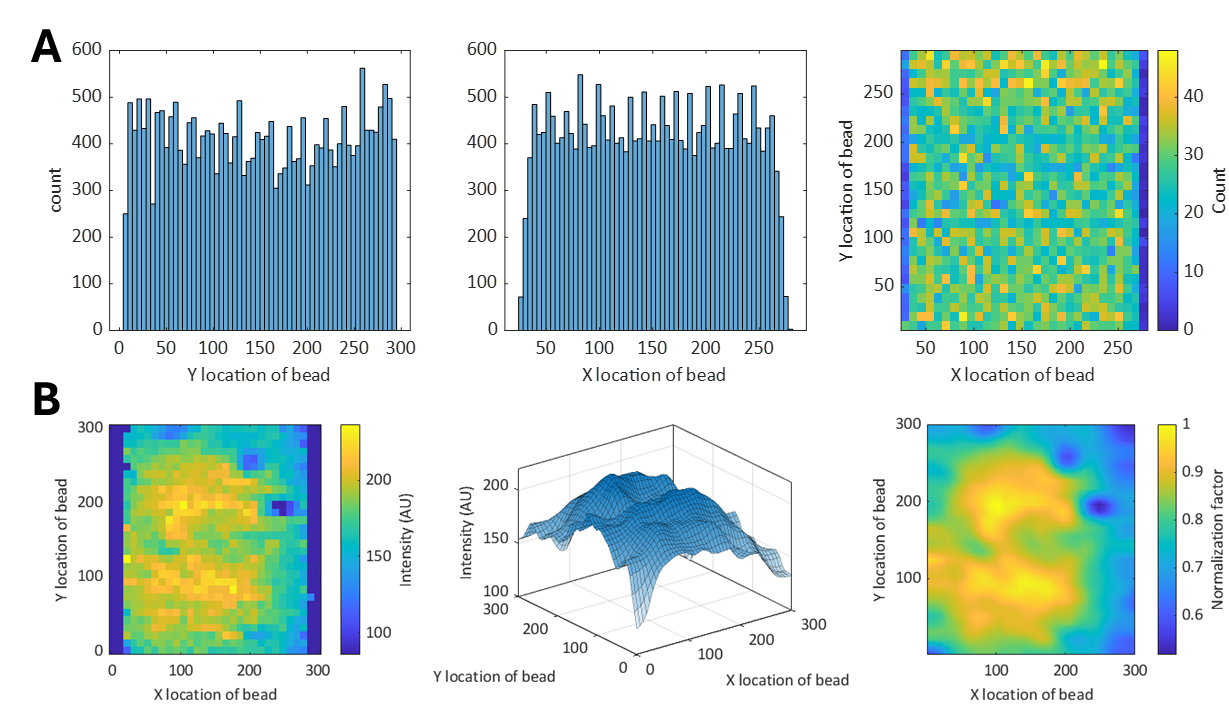

**Figure S8**: Field of view (FOV) characterization for intensity normalization. Diluted beads (not photobleached) were imaged in the system for 24 minutes, resulting in ~24,700 detected localizations. **A.** Bead positions were uniformly distributed across the FOV. Left and middle: histograms of Y and X localization coordinates, respectively. Right: 2D density map of localization positions. **B.** Spatial mapping of fluorescence intensity across the FOV. Left: average bead intensity in each spatial bin. Middle: smoothed surface representation using linear interpolation. Right: normalized intensity map, where each pixel value represents a correction factor. This map was used to normalize fluorescence measurements based on the bead’s position in the FOV, correcting for nonuniform illumination.
